# On Modeling of COVID-19 for the Indian Subcontinent using Polynomial and Supervised Learning Regression

**DOI:** 10.1101/2020.10.14.20212563

**Authors:** Dishita Neve, Honey Patel, Harsh S. Dhiman

## Abstract

COVID-19, a recently declared pandemic by WHO has taken the world by storm causing catastrophic damage to human life. The novel cornonavirus disease was first incepted in the Wuhan city of China on 31^st^ December 2019. The symptoms include fever, cough, fatigue, shortness of breath or breathing difficulties, and loss of smell and taste. Since the devastating phenomenon is essentially a time-series representation, accurate modeling may benefit in identifying the root cause and accelerate the diagnosis. In the current analysis, COVID-19 modeling is done for the Indian subcontinent based on the data collected for the total cases confirmed, daily recovered, daily deaths, total recovered and total deaths. The data is treated with total confirmed cases as the target variable and rest as feature variables. It is observed that Support vector regressions yields accurate results followed by Polynomial regression. Random forest regression results in overfitting followed by poor Bayesian regression due to highly correlated feature variables. Further, in order to examine the effect of neighbouring countries, Pearson correlation matrix is computed to identify geographic cause and effect.

## 1. Introduction

On 31 December 2019, in Wuhan City, Hubei Province of China cases of unknown virus were detected. From 31 December 2019 through 10 January 2020, a total of 44 case-patients of unknown etiology were reported to WHO by the national authorities in China. The authorities in China identified a new type of virus called coronavirus, which was isolated on 7 January 2020. On 11 and 12 January 2020, WHO received detailed information from the National Health Commission of China that the outbreak of virus is originated from the unhygienic wet seafood market in Wuhan City. On 12 January 2020, China shared the genetic sequence of the novel coronavirus. [1]

COVID-19 is an infectious disease caused by severe acute respiratory syndrome coronavirus (SARS-CoV-2). Common symptoms include fever, cough, fatigue, shortness of breath or breathing difficulties, and loss of smell and taste. [2] While most people have mild symptoms, some people develop acute respiratory distress syndrome (ARDS) possibly precipitated by cytokine storm, [3]multi-organ failure, septic shock, and blood clots. The incubation period may range from one to fourteen days. [4]

The first case of COVID-19 in India, which originated from China, was reported on 30 January 2020. India currently has the largest number of confirmed cases in Asia, [5]and has the second-highest number of confirmed cases in the world after the United States, [6] [7] with the number of total confirmed cases breaching the 100,000 mark on 19 May, [8]and 1,000,000 confirmed cases on 17 July 2020. On 29 August 2020, India recorded the global highest single-day spike in COVID-19 cases with 78,761 cases, surpassing the previous record of 77,368 cases recorded in the US on 17 July 2020. [9] India currently holds the single day record for largest increase in cases, set on September 17, with an additional 97,894. [10]

The World Health Organization declares Covid-19 a pandemic, which is defined “an epidemic that has spread over several countries or continents, and most people do not have immunity against it”. At this point, more than 1,21,000 people were infected and over 4,300 died globally. To reduce Corona virus cases, India government introduced 21 days lockdown on 23 March 2020 with several special rules. The lockdown was extended till 31 May 2020 with several changes in guidelines. The Government of India has proposed multiple lockdowns to curb the spread of this virus. Due to these lockdowns, there has been a decrease in the number of cases from 11.8% to 6.3% daily. But the government cannot shut the entire nation forever as the economy may fall drastically. Due to lockdown the country is facing job crisis and unemployment. The GDP has also decreased to –23.9%. The Tourism sector had been highly affected due to lockdown. Also, Textile industry is highly affected. According to World Trade Organization (WTO) and Organization for Economic Cooperation and Development (OECD) have indicated COVID-19 pandemic as the largest threat to global economy since the financial emergency of 2008–2009. This emergency has been the largest emergency till date. So, COVID-19 has undoubtedly put forth a remarkably adverse effect on the day to day life of the entire human society and on world economy.

Medical researchers throughout the globe are currently involved to discover an appropriate vaccine and medications for the disease. On 23 March, the National Task Force for COVID-19 constituted by the ICMR recommended the use of hydroxychloroquine for the treatment of high-risk cases. Since there is no approved medication till now for killing the virus so the governments of all countries are focusing on the precautions which can stop the spread. In India Serum Institute of technology is working on clinical trial to discover corona vaccine.

Machine learning (ML) has proved itself as a prominent field of study over the last decade by solving many very complex and sophisticated real-world problems. These prediction systems can be very helpful in decision making to handle the present scenario to guide early interventions to manage these diseases very effectively. To contribute to the current human crisis our attempt in this study is to analyze COVID-19 spread in India. Hence a verified and proved vaccine of this coronavirus is not found and many researches and scientific experiments are undergoing and some vaccines are still in trial stage so to utilize the available resources and to deal with the current ongoing pandemic, modelling the situation and analyzing the outcome the future can be predicted.

In this study first we studied the growth curve of the current situation. Then different regression models like Polynomial Regression, Forest Regression, Support Vector Regression, Naive Bayes, were used to predict the situation till September 7, 2020 and an optimal model was proposed. This is especially useful for Government and health workers to analyze current situation and predict the situation up to few weeks. This helps them to take necessary precautions and actions to curb the spread of disease. We have divided data set into 4 regions to understand which polynomial curve fits best. We have also compared our results with neighboring countries like Pakistan, Sri Lanka, and Bangladesh for better analysis.

## 2. Methods

In this section, several methods based on machine learning regression and polynomial regression are discussed. The analysis is carried out for pan India cases where the data is acquired from [13-14]. The experiments for regression analysis are carried out on Python platform.

### 2.1 Polynomial Regression Method

Regression models are statistical sets of processes which are used to estimate or predict the target or dependent variable based on dependent variables. Regression may be linear or nonlinear. A model is said to be linear when it is linear in parameters. For example, the second-order polynomial in one variable

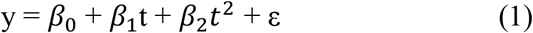

and the second-order polynomial in two variables

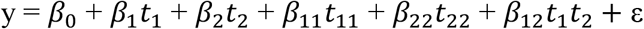

are also the linear model. In fact, they are the second-order polynomials in one and two variables, respectively [2].

Polynomial regression is a special type of regression which works on the curvilinear relationship between the dependent values and independent values. For COVID 19 we have used polynomial regression with only one variable.

Polynomial with one variable

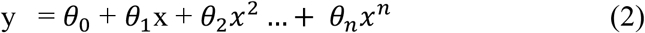

Equation 2 shows the relationship between a dependent and independent variable in polynomial regression. In Equation (2), x is the independent variable and *θ*_0_ is the bias also the intercept and *θ*_1_, *θ*_2_ …. *θ*_*n*_are the weight or partial coefficients assigned to the predictors and n is the degree of polynomial.

The polynomial regression used in the study includes transformation of data into polynomials and applying linear regression to fit the parameter. A polynomial regression with degree equal to 1 is a linear regression. Choosing the value of a degree is a challenging task. If the degree of polynomial is less, it will not be able to fit the model properly and if the value of degree of polynomial is greater than actual, it will overfit the training data.

The mean squared error (MSE) is an unbiased estimator of the variance σ of the random error term and is defined in equation

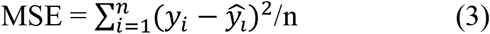

where *y*_*i*_ are observed values and 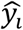 are the fitted values of the dependent variable Y for the ith case. Since the mean squared error is the average squared error, where averaging is done by dividing by the degrees of freedom, MSE is a measure of how well the regression fits the data. The square root of MSE is an estimator of the standard deviation *σ* of the random error term. The root mean squared error 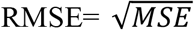 is not an unbiased estimator of σ, but it is still a good estimator. MSE and RMSE are measures of the size of the errors in regression and do not give a indication about the explained component of the regression fit.

### 2.2 Random Forest Regression

Random forest is an ensemble method that generates something akin to a forest of trees from a given training sample. Ensemble-based models are far more accurate than a single method owing to advantages like capturing linearity and nonlinearity of time series obtained from individual methods [11].

A random forest begins with splitting the input features into a group of subsets that essentially form a tree. A particular tree is characterized by a node that leads to a number of branches, as depicted in Figure. 1

**Figure 1:**
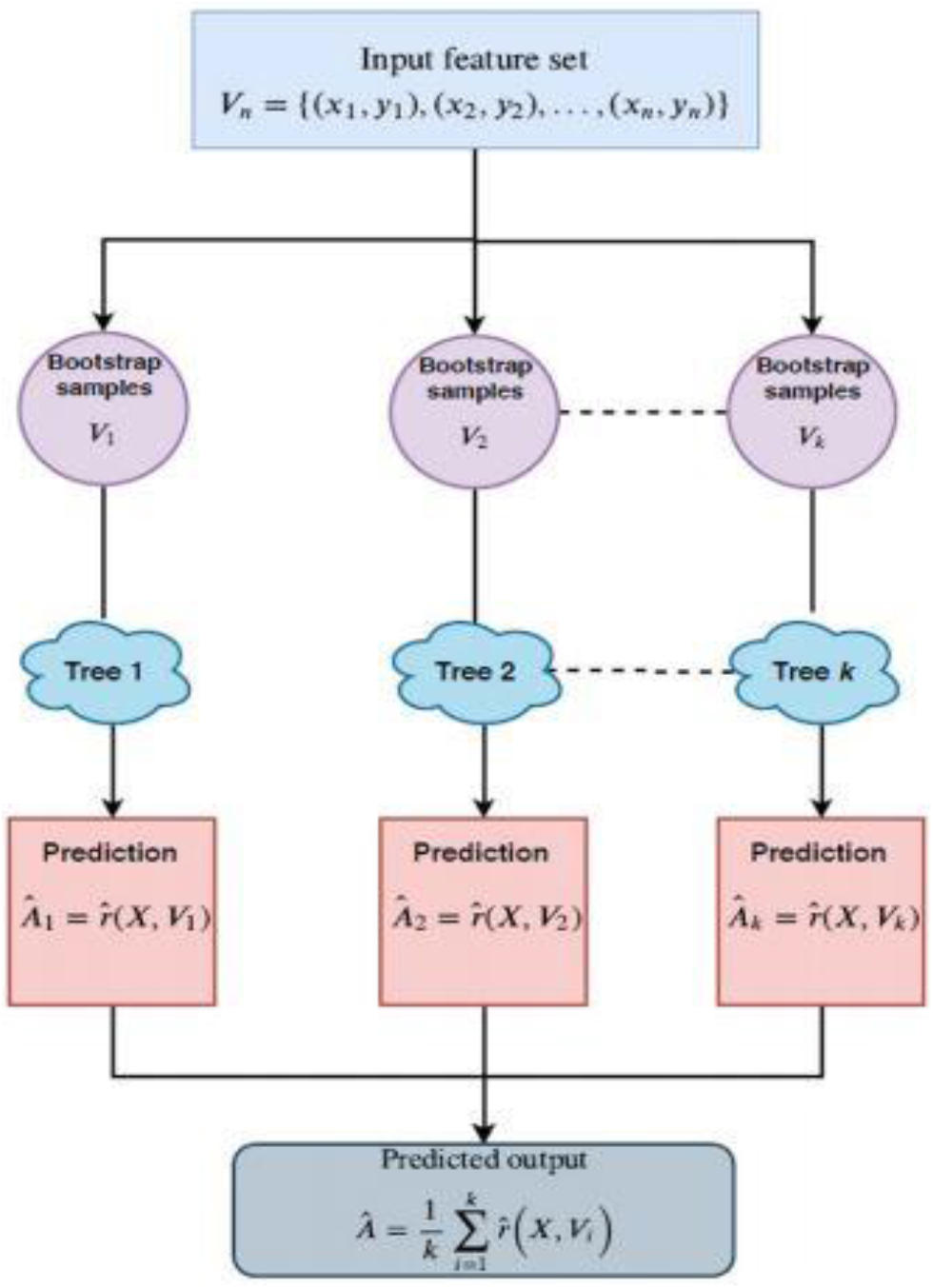
Flowchart of Random forest regression

In random forest regression, the number of trees and the number of random features in each tree decomposition are the parameters that decide the performance of regression. At each decision tree, a fitting function is created, which acts on the random features selected. Finally, at the end of the training process a random forest model is created. It is worth noting that during the training process, each tree is created from randomly selected input vectors and thus is called a “random” forest. Mathematically, the estimated output of a random forest regression is given as

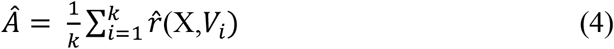

Where 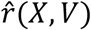 is the representative tree at the end of training process, X is the set of input feature vectors, T is the collective set representing an input–output pair *V*_*i*_ = (*x*_1_,*y*_1_), (*x*_2_,*y*_2_) … (*x*_*k*_,*y*_*k*_) The predicted output is averaged over k decision trees. Decision trees are sensitive to the data on which they are trained. Changing the training data can change the predictions.

Meanwhile a common problem of overfitting persists in machine learning regression models when a well-trained model captures the noise component as well. To reduce the complexities posed by overfitting, a random forest makes a compromise between flexible and inflexible models. In the training phase, each regression tree repeatedly draws a sample from the feature set. This ensures that even though the tree may possess a high variance, the overall variance of the forest may be low. A random forest works on the principle of bagging, which combines the predictions from different tree models to give an overall insight to the data under training. This also helps to reduce the potential overfitting caused by supervised machine learning models.

### 2.3 Support Vector Regression

The main objective of SVM is to find the optimal hyperplane which linearly separates the data points in two component by maximizing the margin [12].

Consider a set of data(*x*_1_,*y*_1_), (*x*_2_,*y*_2_) … (*x*_*k*_,*y*_*k*_) ⊂ X x R, where X denotes the input feature space of dimension *R*^*n*^. Let Y = (*y*_1_, *y*_2_ …. *y*_*n*_) denote the set representing the training output or response, where i = 1, 2,…, n and *y*_*i*_ ∈ R.

This type of SVR uses an ε-insensitive loss function that intuitively accounts for sparsity similar to SVR by ignoring errors less than ε. ε-SVR aims to find the linear regressor

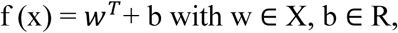

For prediction, where x ∈ X is the input set containing all the features, w is the weight coefficient related to each input xi, and b is the bias term. The objective is to find f (x) with maximum deviation ε from the respective feature sets while being as flat as possible.

In case of COVID – 19 features sets might not be able to linearly separable. To handle such nonlinearities in the feature sets, the so called kernel functions are used to transform data to a higher-dimensional space (“kernel trick”). After transformation via a suitable mapping function φ : Rn → Z, the data become linearly separable in the target space (high-dimensional space), that is, Z. The inner product <*w*^*T*^,φ(x)> in the target space can be represented by using a kernel function. Kernel functions are similarity functions that satisfy Mercer’s theorem such that k(*x*_*i*_, *x*_*j*_) = φ(*x*_*i*_), φ(*x*_*j*_) are the elements of the kernel matrix K. Several kernel functions are available in the literature like linear, polynomial of degree d, Gaussian, and the radial basis function (RBF) with bandwidth of the function σ and exponential function

Fig. 4.2 illustrates the kernel trick used when the input vectors are not linearly separable. This transformation makes the computation of weights and bias vector much easier. Now we look at the dual form of the optimization problem:

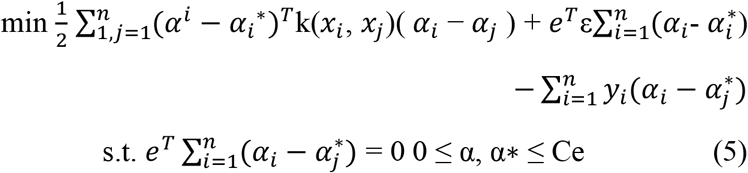

where α and *α*^∗^ represent the positive and negative Lagrange multipliers such that *α*_*i*_*α*^∗^ = 0, i= 1, 2,…, n. The regressor f (x) can be written as

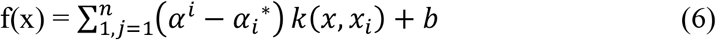

**Figure 2:**
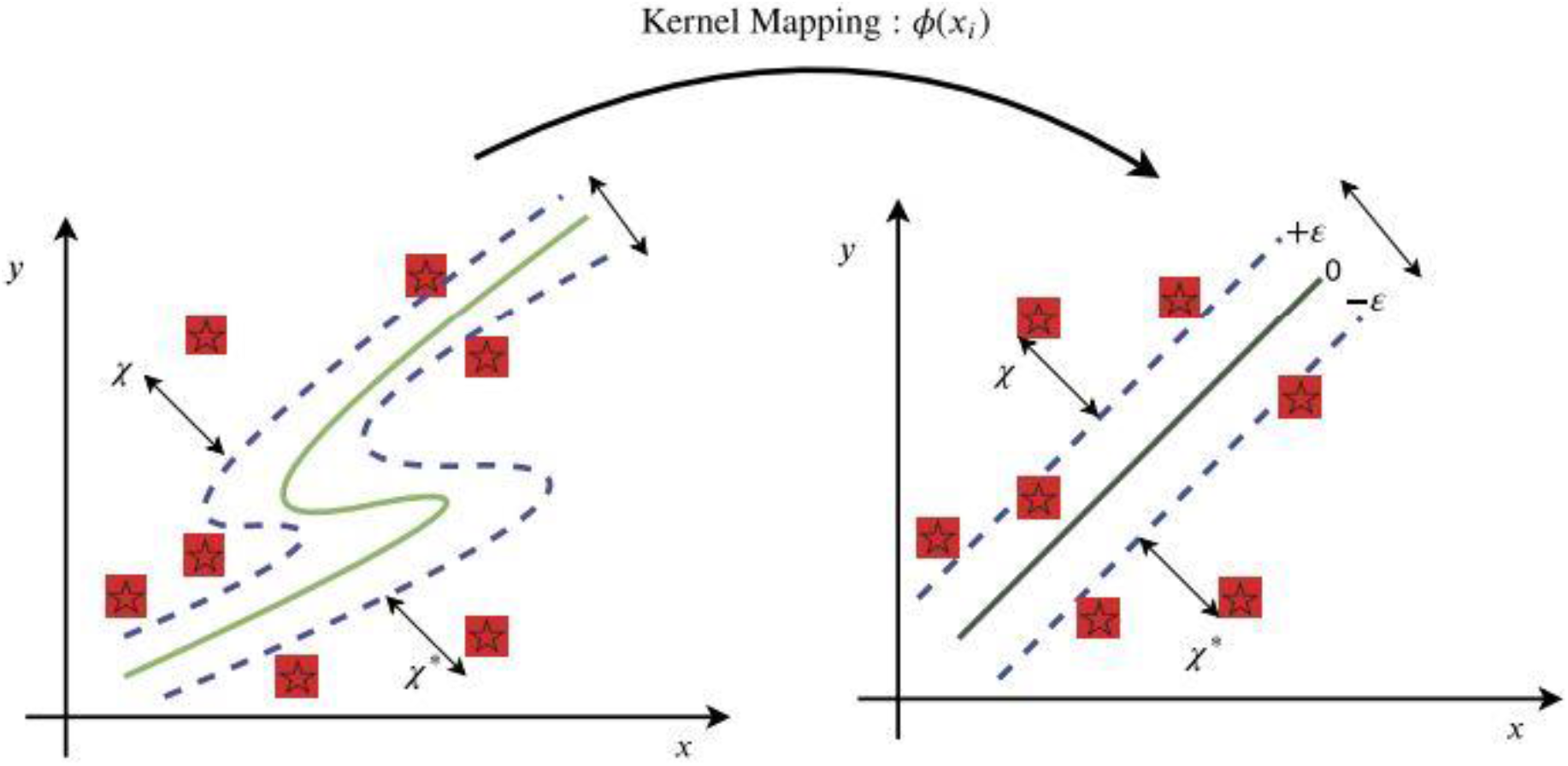
Kernel mapping into higher-dimensional space for nonlinear datasets.

**Figure 3.**
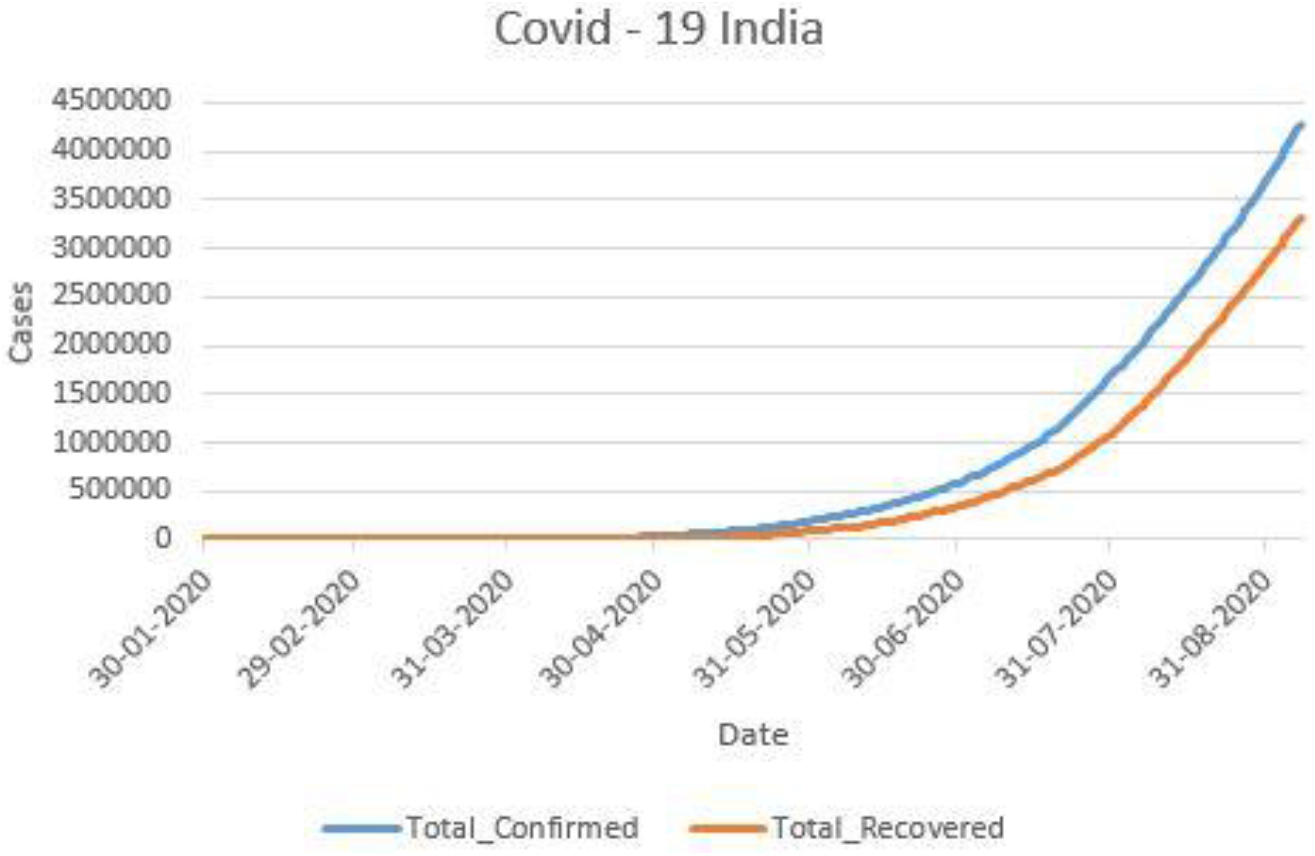
Growth Curve of Total Confirmed Cases and Total Confirmed Cases

**Figure 4.**
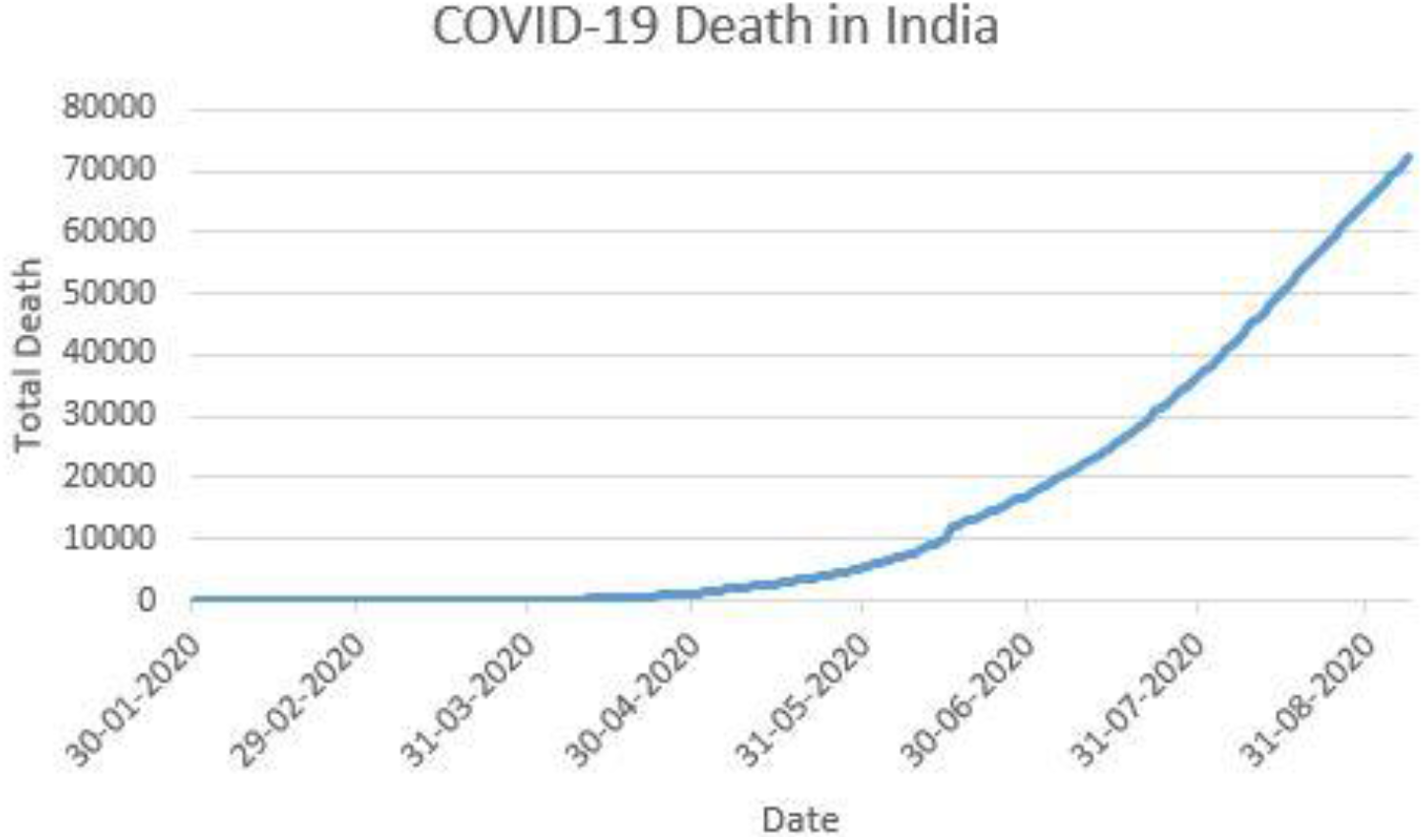
Growth Curve of Deceased Cases

**Figure 5.**
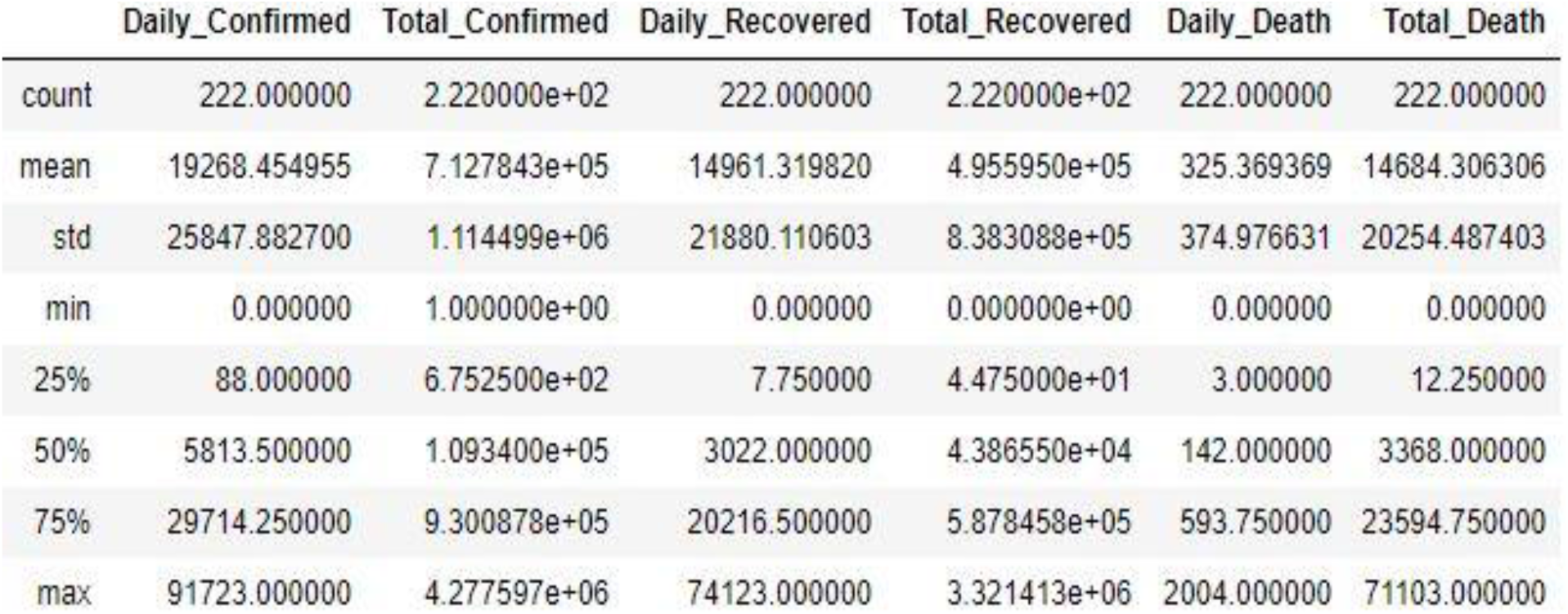
Statistical Analysis of COVID-19 Pandemic in India

**Figure 6.**
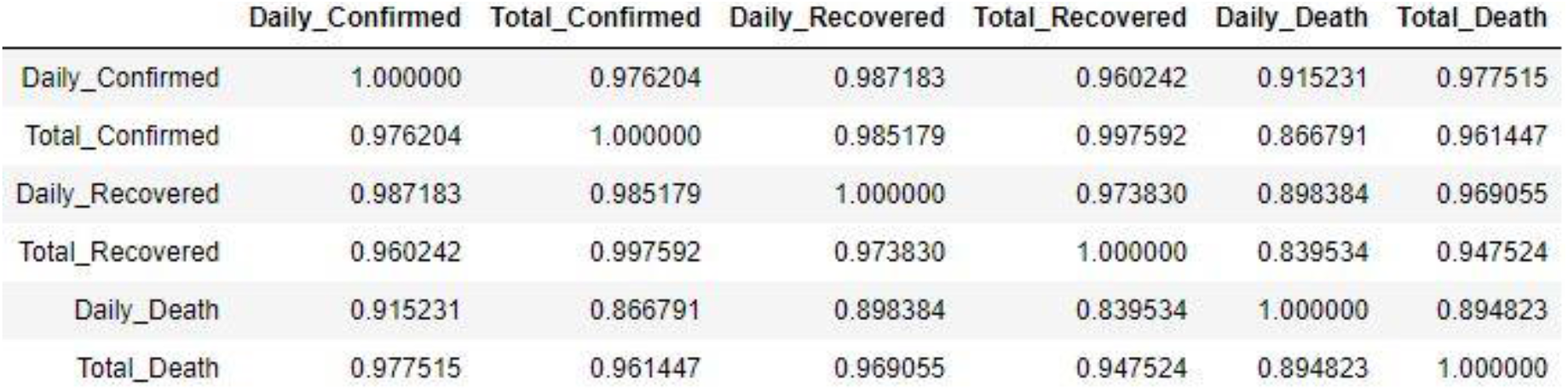
Correlation Analysis of Total Confirmed, and Recovered and Daily Death and Total Death

**Figure 7.**
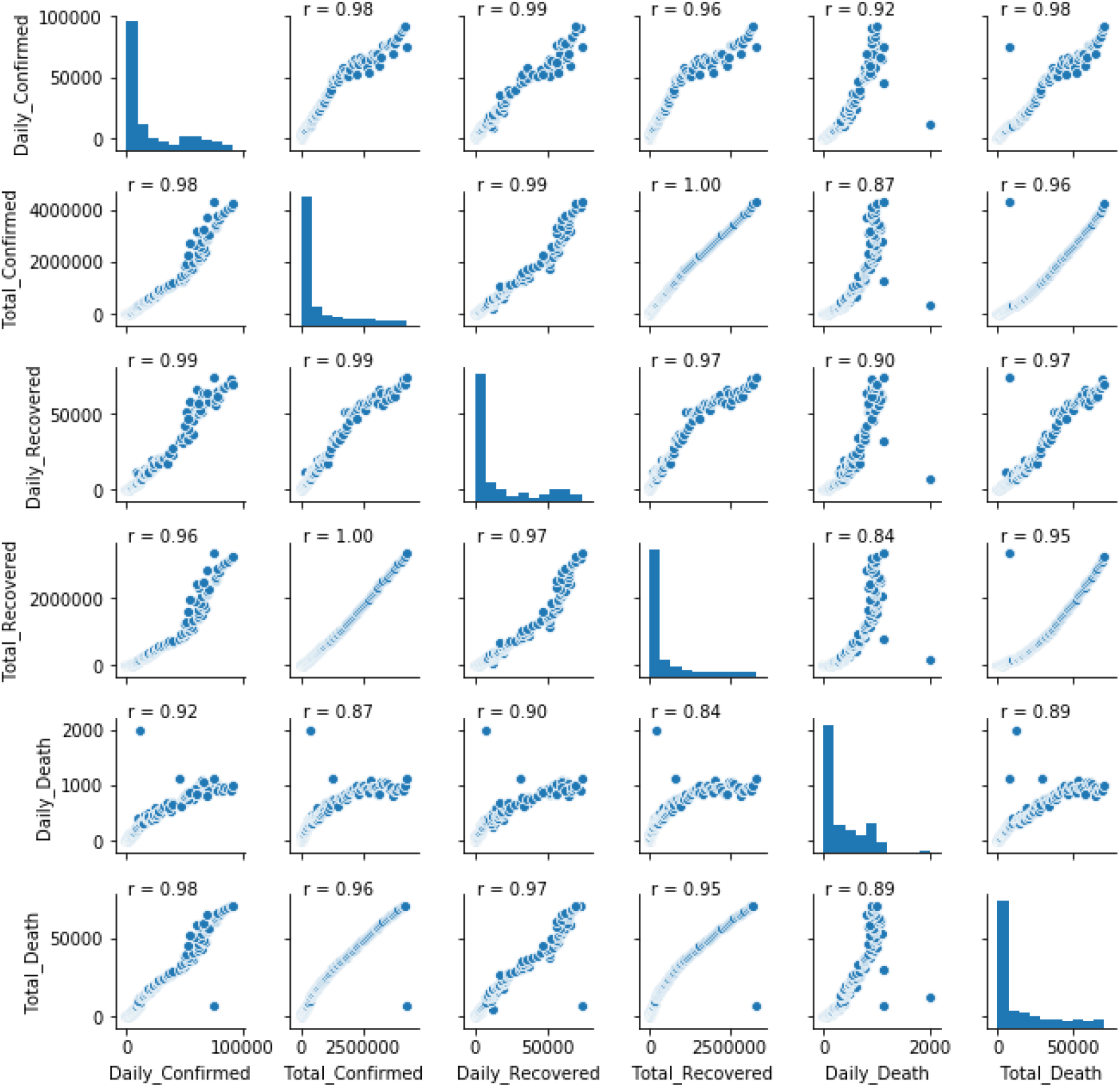
Scatter plot between feature variables

**Figure 8:**
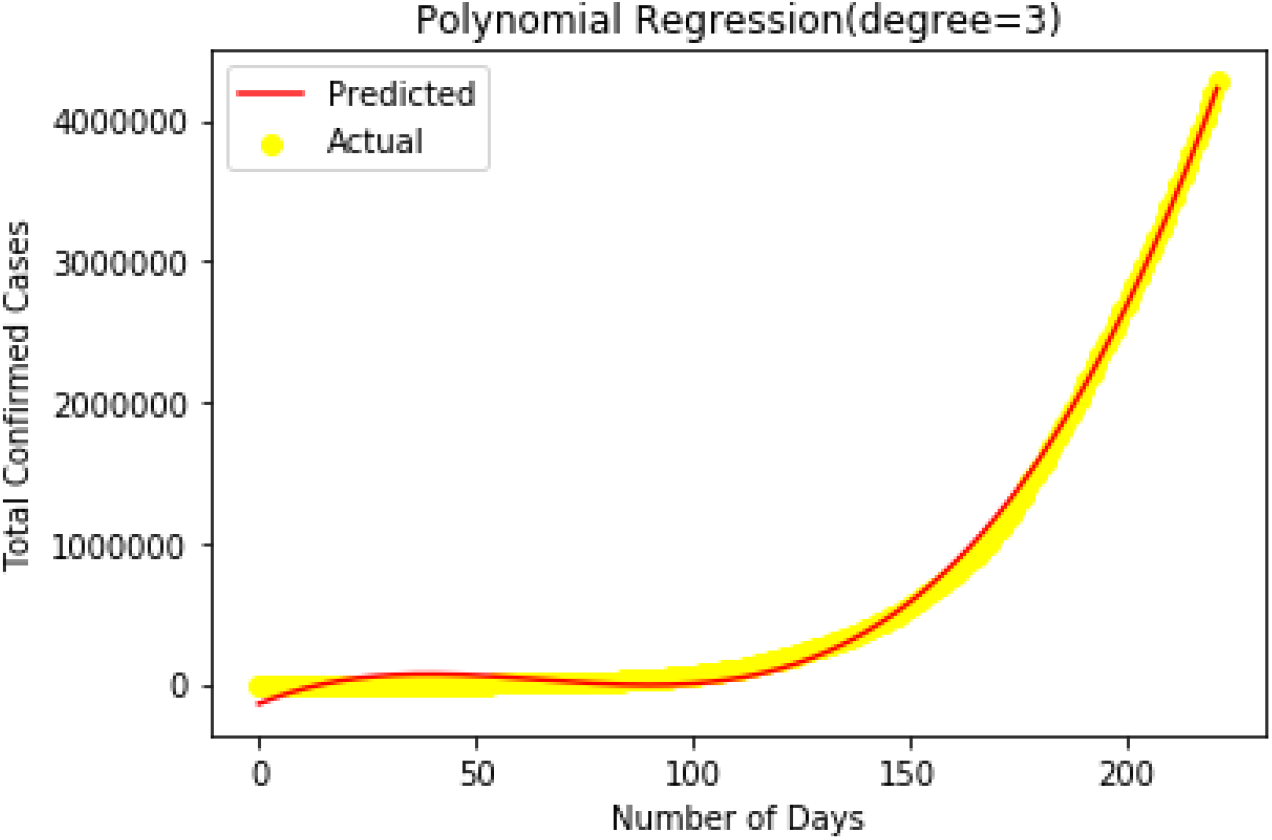
Prediction using polynomial regression

The complexity of this regressor is independent of the dimensionality of the feature set and only depends on the number of support vectors, which are nothing but the data points that separate the feature sets from each other. However, the performance of the SVR also depends on the choice of a kernel function and helps in reducing the computation time of the regression.

### 2.4 Naïve Bayes Regression

Naive Bayes assigns a probability to every possible value in the target range. The resulting distribution is then condensed into a single prediction. In categorical problems, the optimal prediction under zero-one loss is the most likely value—the mode of the underlying distribution. However, in numeric problems the optimal prediction is either the mean or the median, depending on the loss function. These two statistics are far more sensitive to the underlying distribution than the most likely value: they almost always change when the underlying distribution changes, even by a small amount. Therefore, when used for numeric prediction, naive Bayes is more sensitive to inaccurate probability estimates than when it is used for classification

Consider the problem of predicting a numeric target value Y, given an example E. E consists of m attributes X_1_, X_2_, …, X_m_. Each attribute is either numeric, in which case it is treated as a real number, or nominal, in which case it is a set of unordered values.

If the probability density function p(Y |E) of the target value were known for all possible examples E, we could choose Y to minimize the expected prediction error. However, p(Y |E) is usually not known, and has to be estimated from data. Naive Bayes achieves this by applying

Bayes theorem and assuming independence of the attributes X_1_, X_2_, …, X_m_ given the target value Y. Bayes’ theorem states that

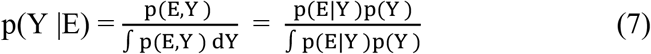

where the likelihood p(E|Y) is the probability density function (pdf) of the example E for a given target value Y, and the prior p(Y) is the pdf of the target value before any examples have been seen. Naive Bayes makes the key assumption that the attributes are independent given the target value, and so Equation 1 can be written

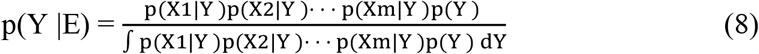

Instead of estimating the pdf p(E|Y), the individual pdfs p(Xi |Y) can now be estimated separately. This dimensionality reduction makes the learning problem much easier. Because the amount of data needed to obtain an accurate estimate increases with the dimensionality of the problem, p(Xi |Y) can be estimated more reliably than p(E|Y).

## 3. Result & Discussion

We have analyzed the data from January 30, 2020 to September 7, 2020. Figure 3 and 4 illustrate that new cases of COVID-19, the rate of Total Confirmed Cases and Recovered was increasing at a slow pace but after April 31, 2020 there was an exponential growth in both Total Confirmed Cases and Recovered Cases. As number of deaths increases up to 72,232. However confirmed cases rise increases 4,277,597 while recovered cases rise 3,321,413 till September 7, 2020.Figure 3 showcase statistical analysis of all variables of COVID-19. From Figure 4, we can see that daily confirmed cases, total confirmed cases, daily recovered, total recovered, daily death and total death are highly dependent on each other.

### Polynomial Regression

We applied polynomial regression to find relationship between total confirmed cases verses number of days. For linear, cubic, quadratic and quartic accuracy was 65.54%, 94.85%, 99.7% and 99.9% respectively. As earlier discussed regarding overfitting in polynomial regression and degree 3 was observed as the best fit polynomial which otherwise would not be observed with degree 4 due to overfitting.

**Table 1.**
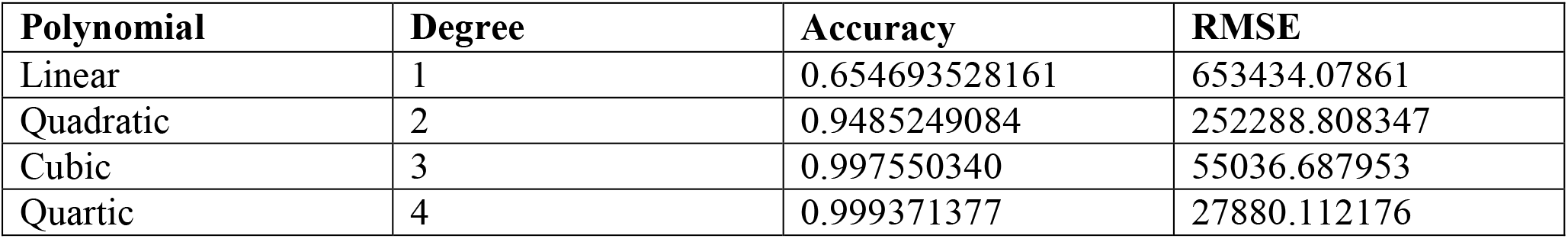
Accuracy and RMSE of Different Degree Polynomial Curves

**Table 2.**
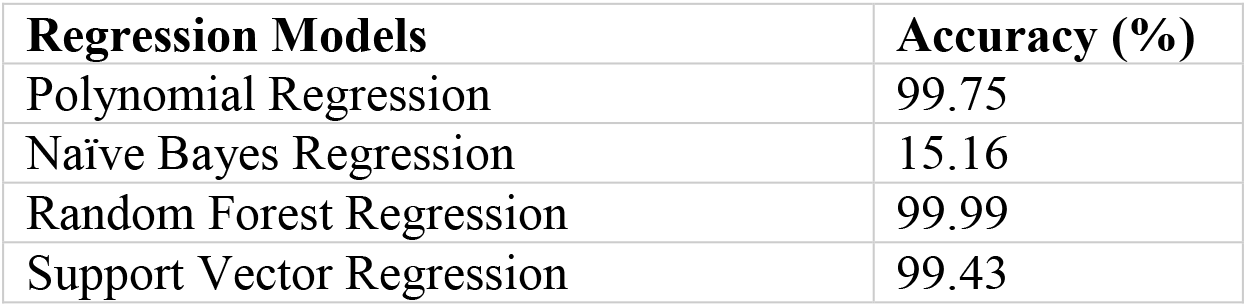
Accuracy of Regression Models

### Naive Bayes Regression

Here we applied Naive Bayes Regression between total confirmed cases and number of days. We get accuracy around 15.56%. From Figure 9 we can see that naïve bayes regression failed to fit the graph and does not show linear relationship.

**Figure 9:**
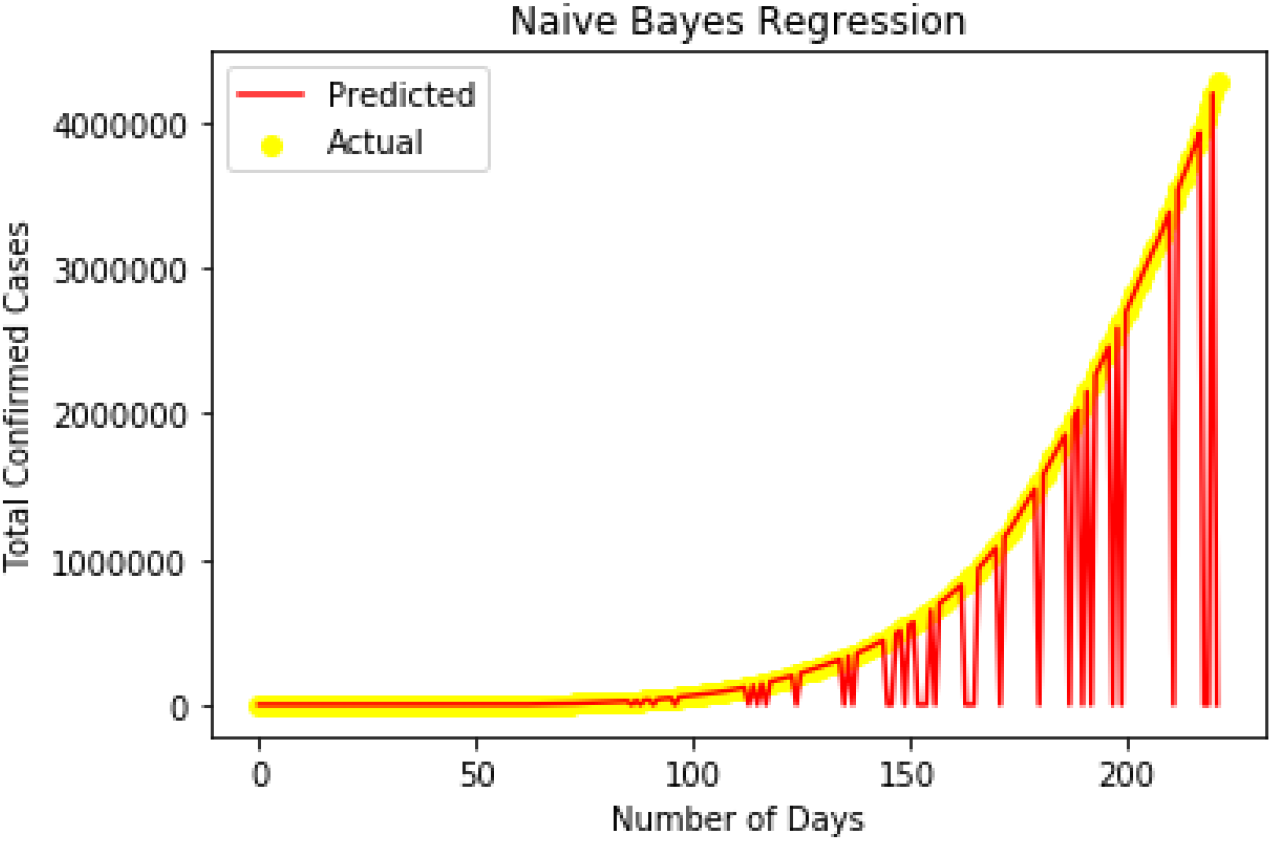
Naive Bayes Regression

### Random Forest Regression

In Figure 10, we have applied Random Forest Regression between total confirmed cases and number of days. Random Forest has successfully fit the growth curve with 99.99% accuracy. But when we predict further value for day 250,275 and 300, we are getting 4221741.4 for all. Technically as number of days increases total confirmed cases cannot be constant so we can observe data overfitting. We apply machine learning models to data set for predicting future values. Random forest failed for future predictions due to overfitting.

**Figure 10:**
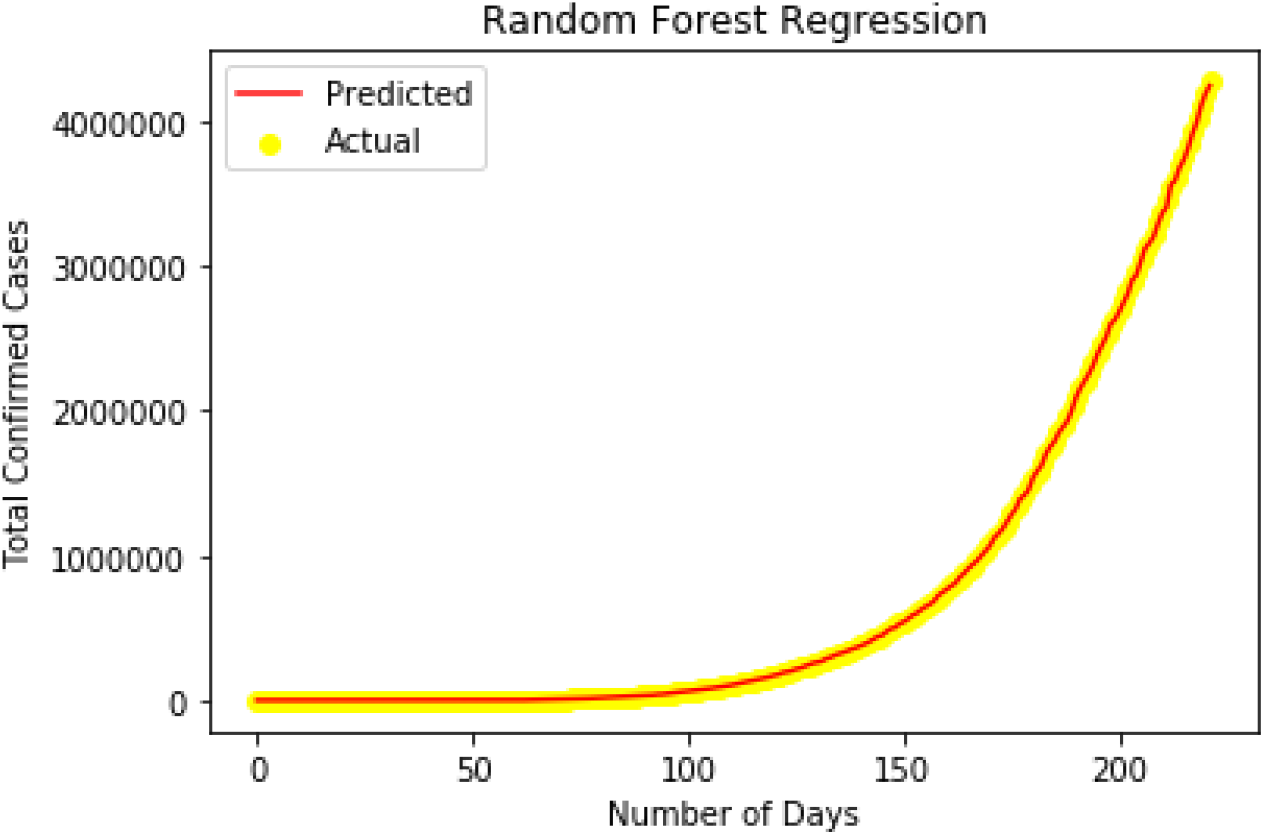
Random Forest Regression

### Support Vector Regression

In Figure 11, we converted number of days and total confirmed cases in standard scalar form for performing better support vector regression. By applying Radial basis function (rbf) kernel we got an accuracy of 99.43%.

**Figure 11:**
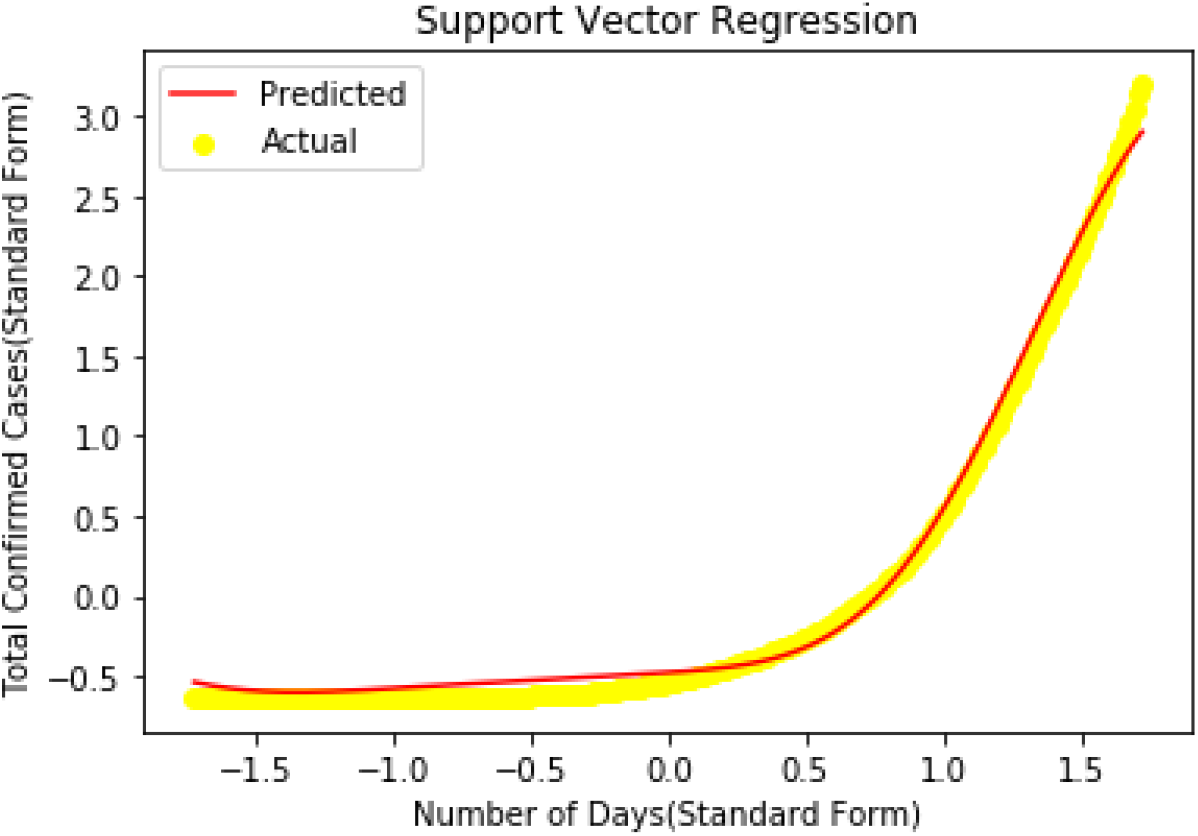
Support Vector Regression

### Polynomial Regression divided in 4 regions

For better understanding among total confirmed cases and number of days we divided total number of days in 4 regions. First region ranges from 30 January 2020 to 24 March 2020. Second region ranges from 25 March 2020 to 18 May 2020. Third region ranges from 19 May to 12 July. Fourth region ranges from 13 July 2020 to 7 September 2020.

For Figure 12, the regression analysis of each region is illustrated in subsequent figures. First region consists of 55 samples from 30 January 2020 to 24 March 2020. In beginning cases increases exponentially of order 5. Accuracy of model is found to be 99.12%. Figure 14 illustrates quadratic curve with an accuracy of 99.37%. In second region, degree of polynomial decreases from 5 to 2. Now, if we increase value of degree predicted curve will overfit the data. Further for region 3 and 4 they show quadratic curve with 99.829% and 99.98%.

**Figure 12.**
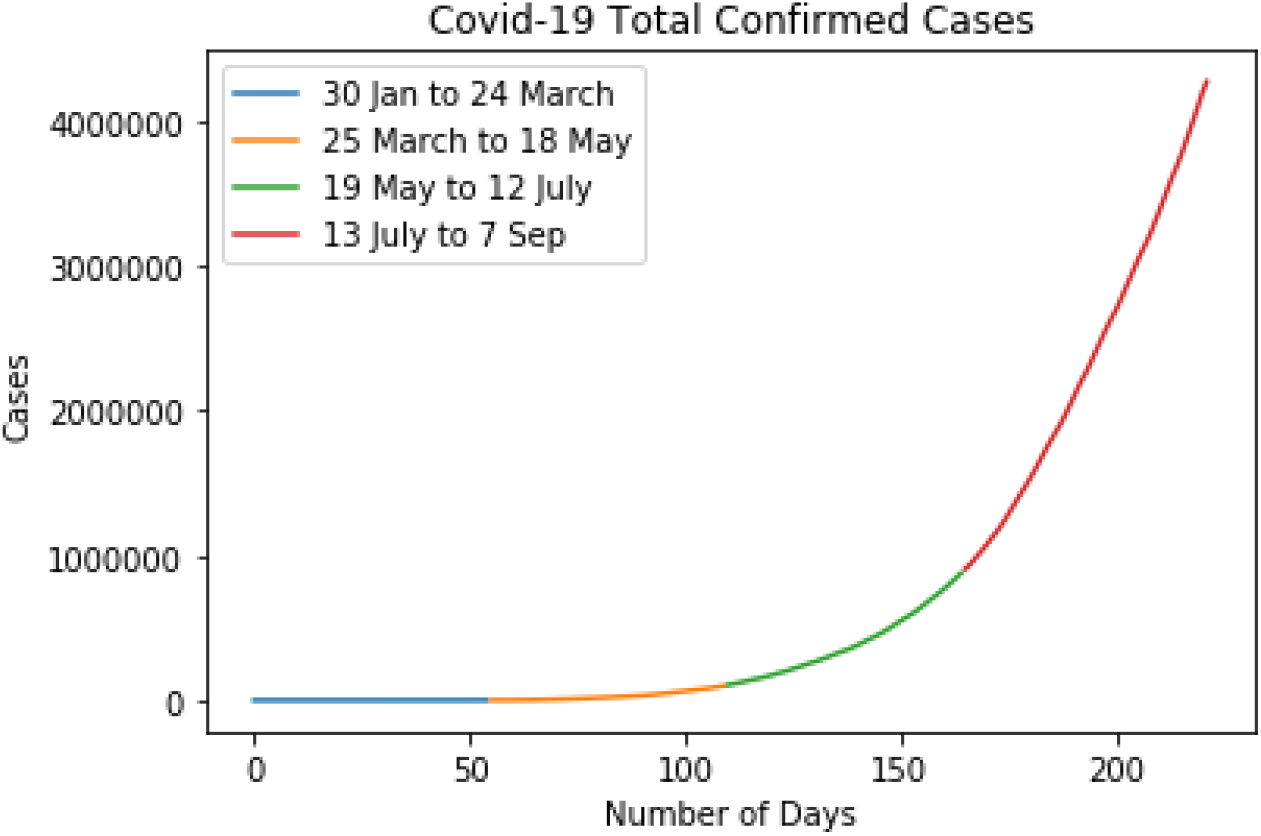
Total Confirmed Cases Divided into 4 regions

**Figure 13.**
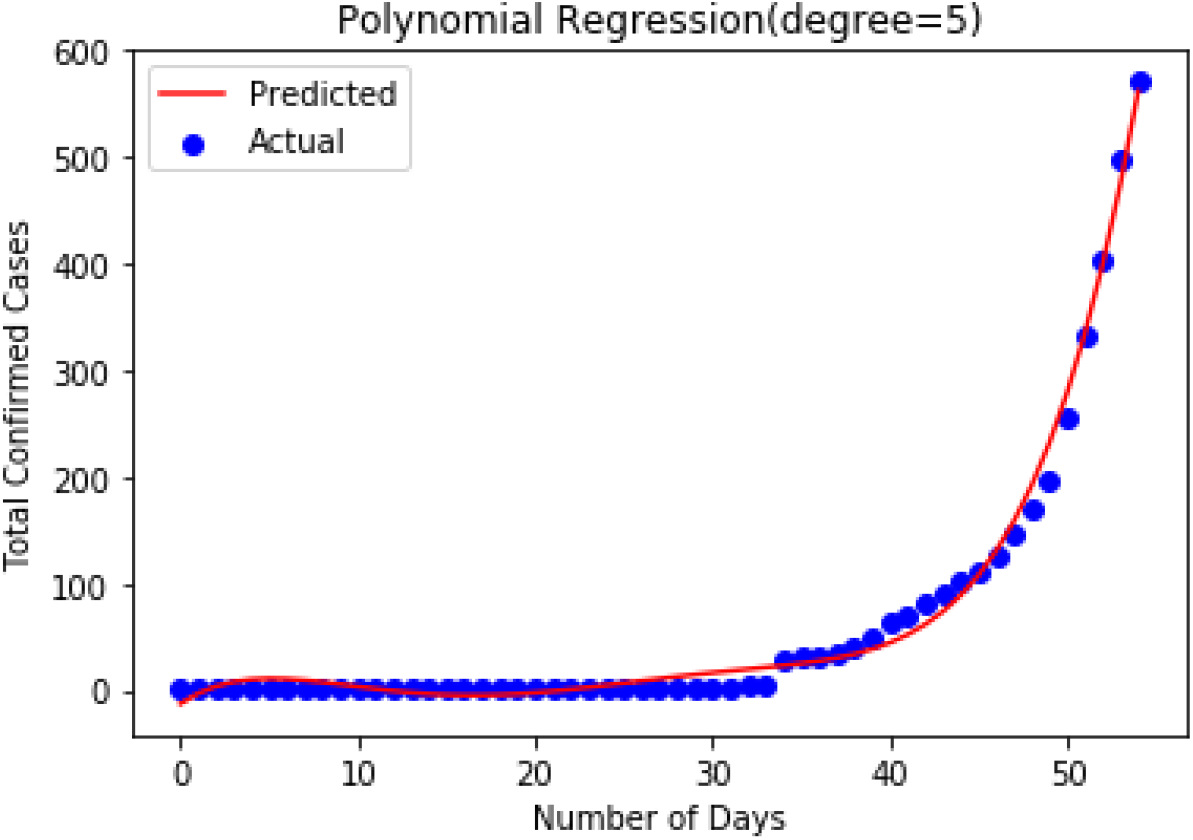
Polynomial Regression for 30 Jan to 24 March

**Figure 14.**
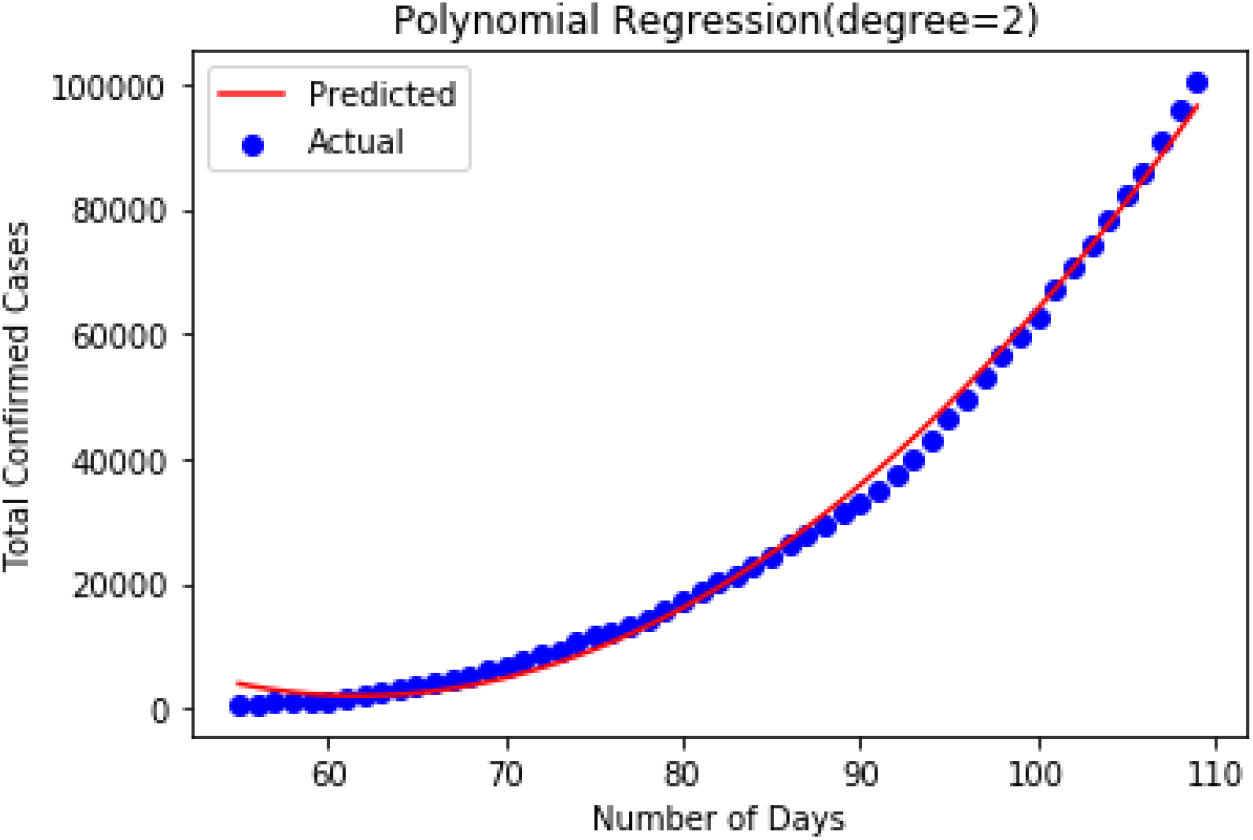
Polynomial Regression for 25 March to 18 May

**Figure 15.**
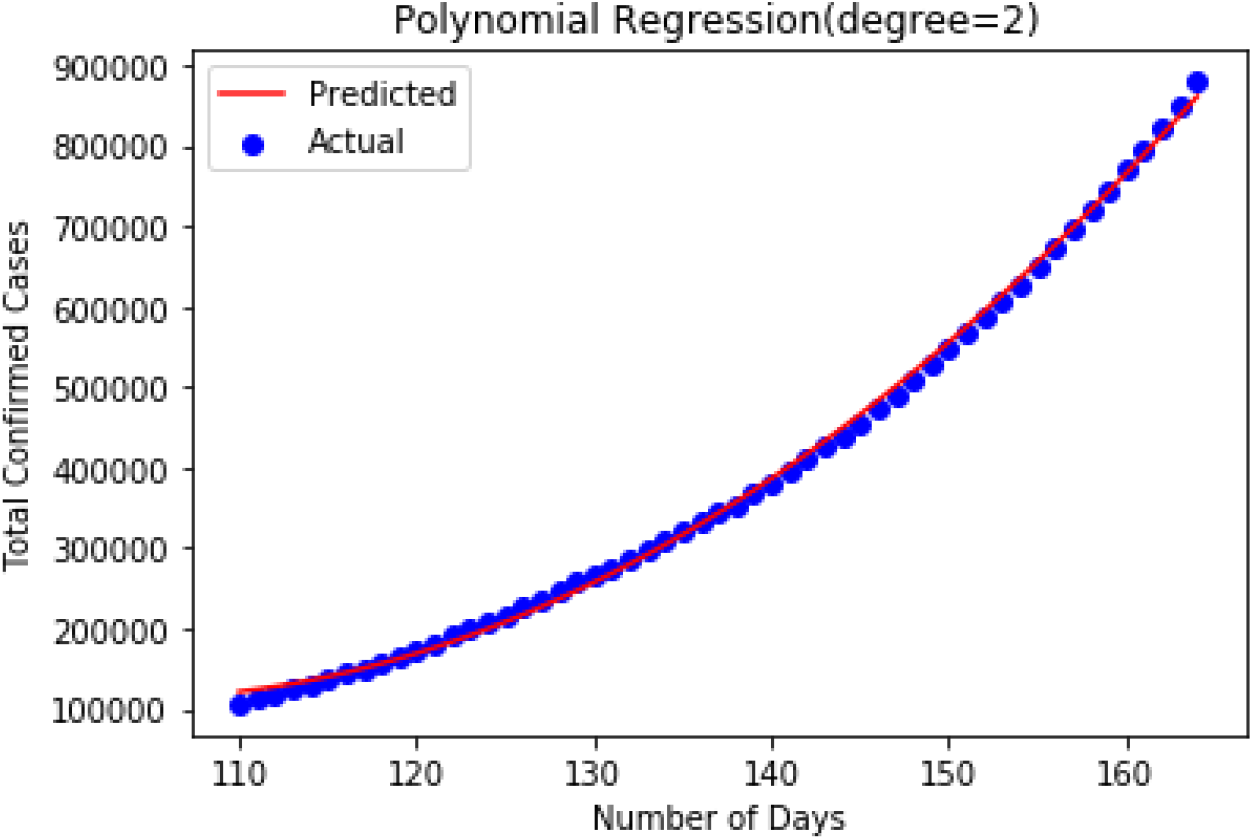
Polynomial Regression for 19 May to 12 July

**Figure 16.**
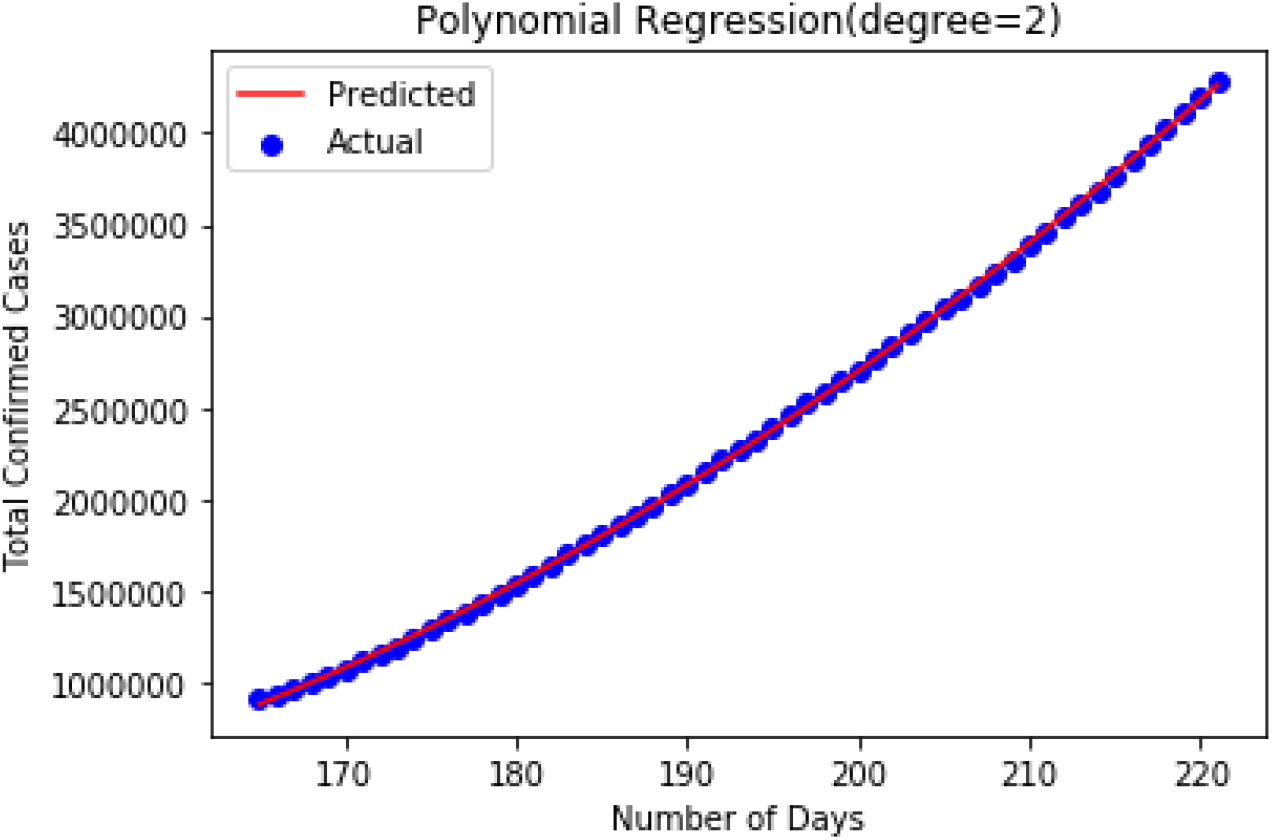
Polynomial Regression for 13 July to 7 Sep

**Table 3:** Performance metrics for subsets using polynomial regression

| Date | Total Days | Degree of Polynomial Regression | Accuracy (%) |
| --- | --- | --- | --- |
| 30 January to 24 March | 55 | 5 | 99.124 |
| 25 March to 18 July | 55 | 2 | 99.37 |
| 19 May to 12 July | 55 | 2 | 99.829 |
| 13 July to 7 September | 57 | 2 | 99.98 |

### Correlation between India, Bangladesh, Sri Lanka and Pakistan (Subcontinent study)

For all three cases total confirmed, total recovered and total death value Pearson coefficient values are majorly dependent on each other. The output data frame can be interpreted as for any cell, row variable correlation with the column variable is the value of the cell. We have calculated the pairwise correlation of all columns in the data frame.

Figure 17-19 illustrate the Pearson correlation matrix between India and neighbouring countries. It is observed that for total confirmed cases, a high correlation coefficient is observed between India and Bangladesh compared to Pakistan and Sri Lanka. Similarly, the correlation coefficient between India and Neighbouring countries is observed to be high and indicates a strong geographic influence in COVID-19 transmission.

**Figure 17.**
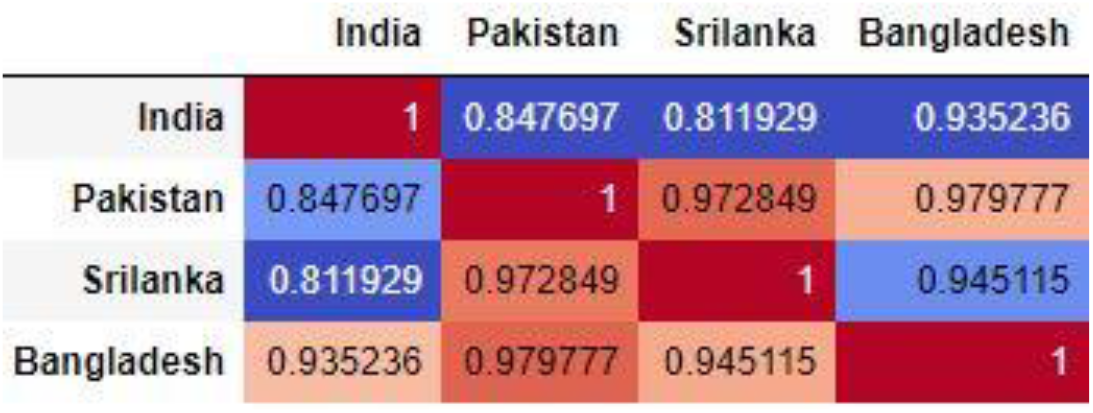
Comparison of Total Confirmed Cases with Neighbouring Countries

**Figure 18.**
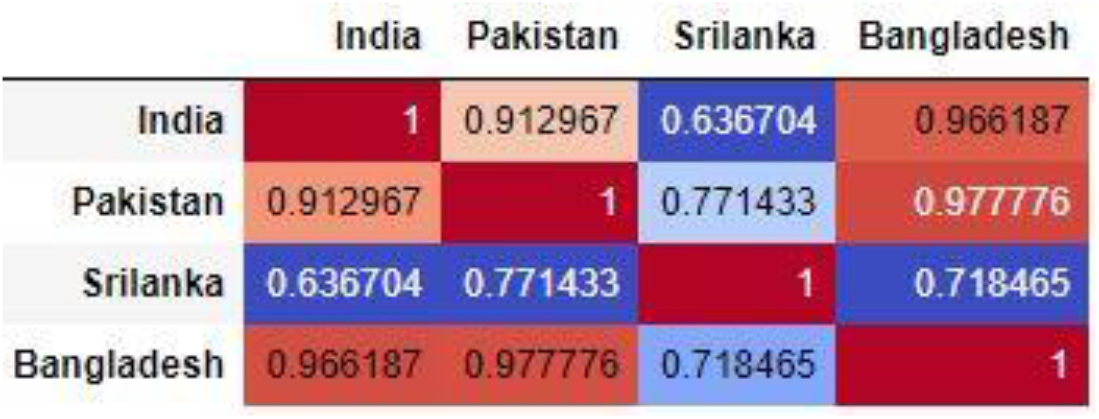
Comparison of Total Recovered Cases with Neighbouring Countries

**Figure 19.**
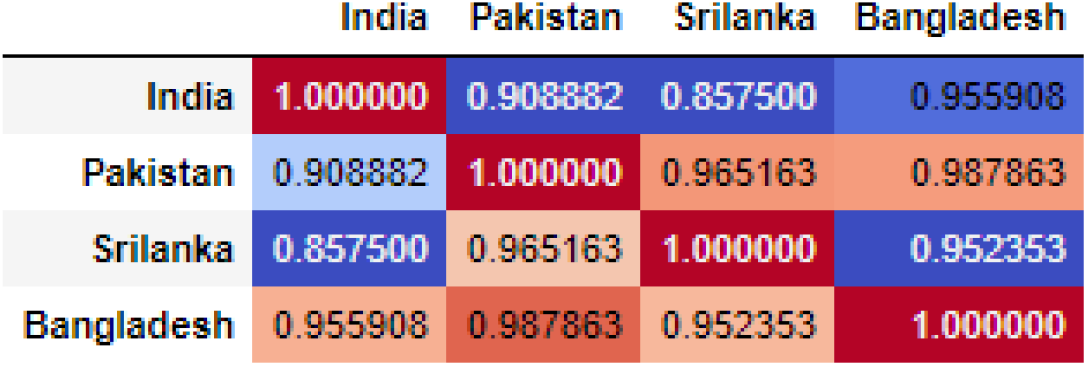
Comparison of Total Death Cases with Neighbouring Countries

## 4. Conclusion

COVID-19 (Coronavirus) has affected day to day life and is slowing down the global economy. This pandemic has affected thousands of peoples, who are either sick or are being killed due to the spread of this disease. Regression analysis is a statistical tool for the investigation of relationships between variables. By applying various regression models from January 30, 2020 to September 7, 2020 we found that Polynomial Regression and Support Vector Regression have performed exceptionally well. But in case of Polynomial Regression, the curve witnessed exponential increase which is not fails to explain the real-time scenario as the total number of confirmed cases will cease in magnitude with improved medication. Naive Bayes regression failed due to less accuracy and Random Forest ended up overfitting the data set. In case of Support Vector Regression with radial basis function kernel, graph decreases after achieving its peak value. So, we can conclude that support vector regression is best among all methods we applied. In beginning COVID 19 cases grow at order of 5 from 30 Jan 2020 to 24 March. From March 25, 2020 to May 18, 2020 COVID-19 cases has increases in order of 2 due to imposition of lockdown. Strict rules were followed during this period. Similar curve has been seen from May 19, 2020 to September 7, 2020.

As the lockdown has uplifted and the life had come to normalcy and as the Government has given authority to open multiplex and cinema halls and slowly and steadily soon the educational institutes will also reopen so, there will be a possibility in sudden spike in the number of cases. Compared to neighboring countries like Bangladesh, Nepal and Pakistan India is worst affected with COVID –19. However, this is due to India’s high population density, increase in testing, large economy activities there is surge in corona virus cases. As Sri Lanka is away from India geographically it is comparatively less affected. We also observe that India and Bangladesh are highly correlated in terms of total confirmed, total recovered and total death. This study is limited to cases because if any government imposes special rule or restriction on international flights than scenario might be different. This study was limited to data-driven models using the total COVID-19 cases. In the future studies, the other co-factors (associated with the demographics, social, cultural, and medical infrastructure, etc.) can be taken to considerations.

## Data Availability

Data can be made available upon reasonable request.

